# Performance of a machine learning model in recognition of normal tissue from histological sections

**DOI:** 10.1101/2021.07.20.21260573

**Authors:** Parikshit Sanyal, Sayak Paul, Avinash Das

## Abstract

**Introduction:** Machine learning and artificial intelligence (AI) models have been applied in histopathology to solve specific problems like detection of metastasis in lymph nodes and immunohistochemical scoring. We have aimed to develop a machine learning model which can be trained in histopathology from the basics, i.e. identification of normal tissue. We have tried to replicate the process through which a human pathologist learns recognition of normal tissue from histological sections, and evaluate the performance of a machine learning model at this task.

**Materials and methods:** A total of 658 histologic images were anonymised, microphotographed at 10x magnification, under the same condition of illumination, with a Magnus DC5 integrated microphotography system. The images were split into two subsets, training (386) and validation (272 images). The images belonged to seven classes of tissue: brain, intestine, kidney, liver, lungs, muscle and skin. Archived material of the hospital were used for the study. A machine learning model using convolutional neural network (CNN) was developed on the Keras platform, using the convolution layers of a pretrained VGG16 model. The model was trained with the training set of images over 10 epochs. After training, performance of the model was assessed on the validation set.

**Results:** The model achieved 88.24% accuracy in classifying the images of the validation set. The most frequent errors were met in recognising images of kidney (14 errors, 33.33%). The commonest error was wrongly classifying kidney tissue as liver (07 errors). Analysis of the deeper layers of the neural network revealed specific patterns in images which were wrongly classified.

**Conclusion:** The results of the present study indicates that a convolutional neural network might be trained in histology similar to a trainee pathologist. The study represents the first step towards developing a machine learning model as a generalised histopathological image classifier.

## Introduction

Histopathology is held as the final bastion of pathology and the gold standard of diagnostic testing. However, interpretation of histopathological sections is inherently observer dependent, and years of training are required to obtain expertise in histopathology. The initial training in histopathology is spent in long hours looking at sections of normal tissue, and preparing a generalised mental representation of each kind of tissue. The trainee pathologist must recognise and classify normal tissue, and this comprises a significant proportion of his/ her training in pathology.

The identification of tissues from histological sections has long been held to be the domain of professionally trained human beings. However, present day machine learning models have matured sufficiently enough, so much so that the task of identification of histological sections by machines can be taken up.

Machine learning models have successfully classified real world images from a very large dataset. ^[1]^ Such models have been able to classify images into multiple classes – i.e. animals, cars, people, street signs, objects of daily use. The basis for such models is usually a artificial neural network (ANN).^[2]^ ANNs are a departure from conventional image analysis techniques. An ANN is constructed of multiple feed forward layers of simpler units (‘neurones’), each of which take an input number *x* and convert it to a non linear output

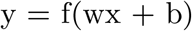

Where ‘w’ and ‘b’ are the two parameters ‘weight’ and ‘bias’ of the particular neurone and f is a non-linear function. A neural network is made of several such layers made of individual neurones. Over repeated epohcs of training, the network adjusts its parameters so as to produce a correct result majority of the time.

For image recognition, a specialised class of ANNs, the convolutional neural network (CNN), is widely in use; the theory of convolutional neural networks has been described in detail by Karpathy et al.^[3]^ CNNs have successfully been applied in several fields of histopathology, such as classification of histopathologic patterns of lung adenocarcinoma^[4]^, for scoring immunohistochemical staining on breast cancers^[5]^, for Gleason’s scoring on prostate cancer^[6] [7]^, mitotic count in breast cancer^[8]^, tumor proliferation, budding and lymphatic vessel density^[9] [10]^, detecting metastasis in lymph nodes^[11]^ and gland segmentation^[12]^.

Such studies have achieved varying levels of success in solving a particular problem. We have taken the systematic approach of training a machine learning model from the ground up, similar to the manner the medical student is trained. Our aim was to develop and test a CNN which can classify normal histological images into seven categories: brain, lung, skin, liver, kidney, muscle, intestine. A similar study has been attempted by Kieffer et al on grayscale images, achieving 74.87% accuracy^[13]^ on 24 classes, using the feature vector from the last layer of a pretrained neural network.

## Materials and methods

We photographed histopathologic slides from 07 classes of tissues: brain, intestine, kidney, liver, lungs, muscle and skin. The histologic sections were retrieved from the archives of the laboratory. All sections were earlier stained with hematoxylin and eosin (staining method is described in Table 1).

**Table 1:** Staining protocol

| Step | Description |
| --- | --- |
| 1 | Sections brought down to water |
| 2 | Placed in haemalum solution for 10 minutes |
| 3 | Rinsed in tap water for 5 minutes |
| 4 | Differentiated in 1% acid alcohol with 3 dips |
| 5 | Washed in running water |
| 6 | Dipped in Scott's solution 3 times |
| 7 | Wash in running water for 3 minutes |
| 8 | Stained in eosin or 15 seconds |
| 9 | Dehydrated and mounted in DPX |

**Table 2:** Distribution of images in training and validation set

| Tissue | Train | Validation |
| --- | --- | --- |
| lungs | 60 | 50 |
| liver | 60 | 42 |
| kidney | 60 | 42 |
| muscle | 58 | 36 |
| intestine | 52 | 45 |
| brain | 52 | 35 |
| skin | 44 | 22 |
|  | 386 | 272 |

**Table 3:** Performance of the model on the validation set

| Predicted | brain | intestine | kidney | liver | lungs | muscle | skin | Total | Accuracy |
| --- | --- | --- | --- | --- | --- | --- | --- | --- | --- |
| Actual |  |  |  |  |  |  |  |  |  |
| brain | 35 | 0 | 0 | 0 | 0 | 0 | 0 | 35 | 100.00% |
| intestine | 0 | 40 | 0 | 0 | 4 | 1 | 0 | 45 | 88.89% |
| kidney | 0 | 0 | 28 | 7 | 3 | 4 | 0 | 42 | 66.67% |
| liver | 2 | 0 | 0 | 39 | 0 | 1 | 0 | 42 | 92.86% |
| lungs | 0 | 0 | 0 | 0 | 50 | 0 | 0 | 50 | 100.00% |
| muscle | 0 | 1 | 0 | 2 | 1 | 31 | 1 | 36 | 86.11% |
| skin | 0 | 4 | 0 | 0 | 1 | 0 | 17 | 22 | 77.27% |
| **Total** | 37 | 45 | 28 | 48 | 59 | 37 | 18 | 272 | 88.24% |

A total of 658 foci were microphotographed, at 10x magnification, with 0.25 numerical aperture, under the same condition of illumination, with a Magnus DC5 integrated microphotography system. The images were split into two subsets, training (386) and validation (272 images). The distribution of images in two datasets is shown as follows.

After collection of images, a machine learning model was developed with the Python^[14]^ programming language and Keras deep learning library; the model constituted of a pretrained image recognition model, VGG16, with the fully connected layers modified to suit the classification problem.^[2] [15]^ The final model consisted of 26 layers of neurones and 2,626,055 trainable parameters (i.e. weights and biases). It accepted a color image of 256 x 192 pixels as input, and produced a single number between 0 to 6 as output (corresponding to the seven classes of images).

The model was trained in the Google Colab platform^[16]^ with 10 epochs, i.e. each training image was shown to the model 10 times. Images were resized to a dimension of 256 x 192 pixels before training. During training, the model adjusted its parameters to minimise the error rate (loss function) at each epoch.

After completion of training, the performance of the model was assessed over the validation set.

## Results

The results are depicted as follows.

The maximum accuracy was seen in recognising the classes ‘brain’ and ‘liver’. Figure 1-4 shows a few images correctly identified by the model.

**Figure 1:**
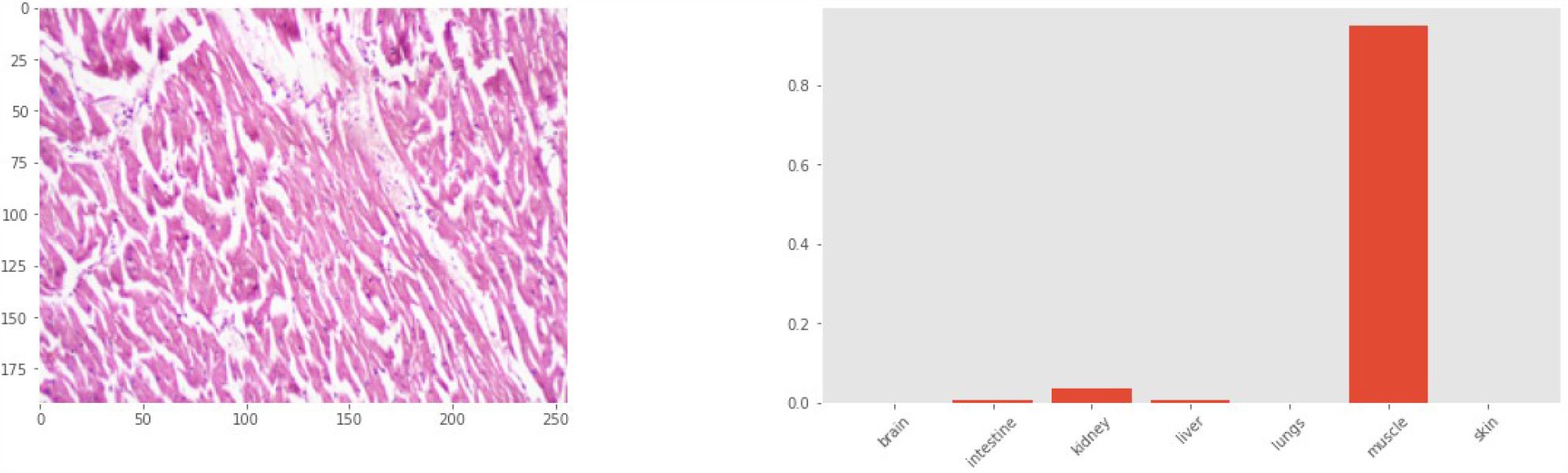
Muscle tissue correctly identified by the model

**Figure 2:**
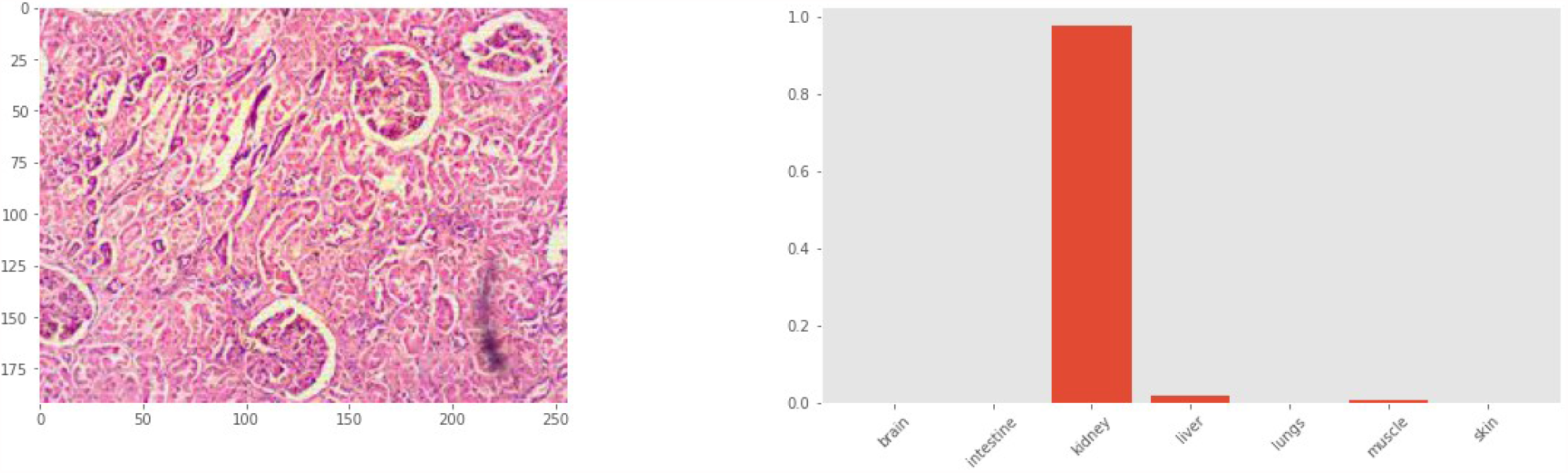
Kidney correctly identified by the model

**Figure 3:**
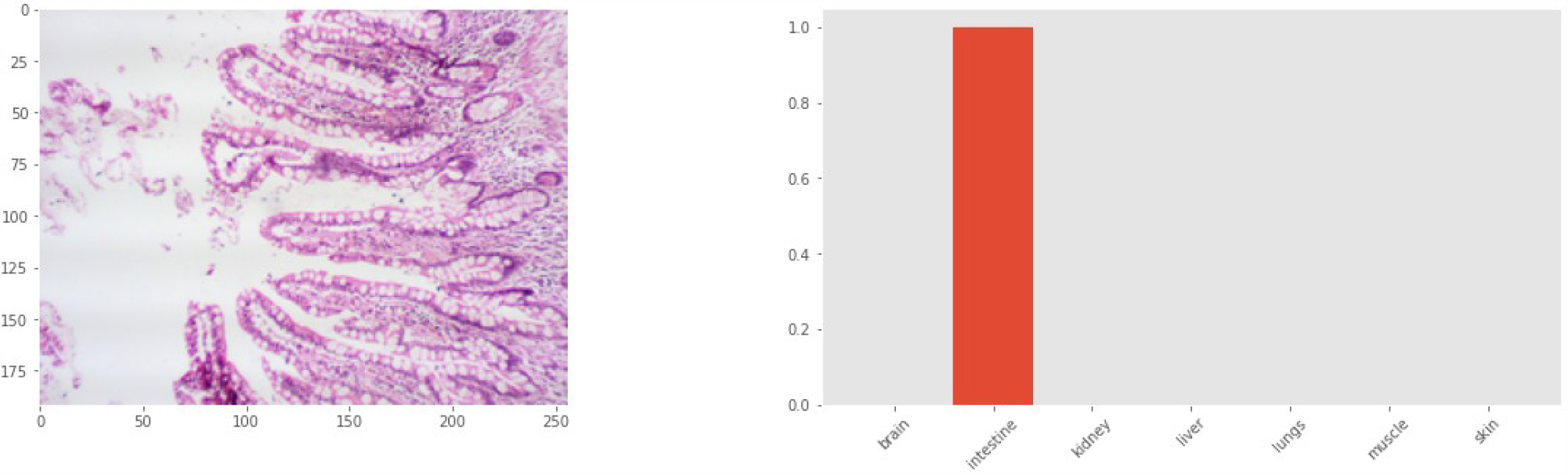
Intestine correctly identified by the model

**Figure 4:**
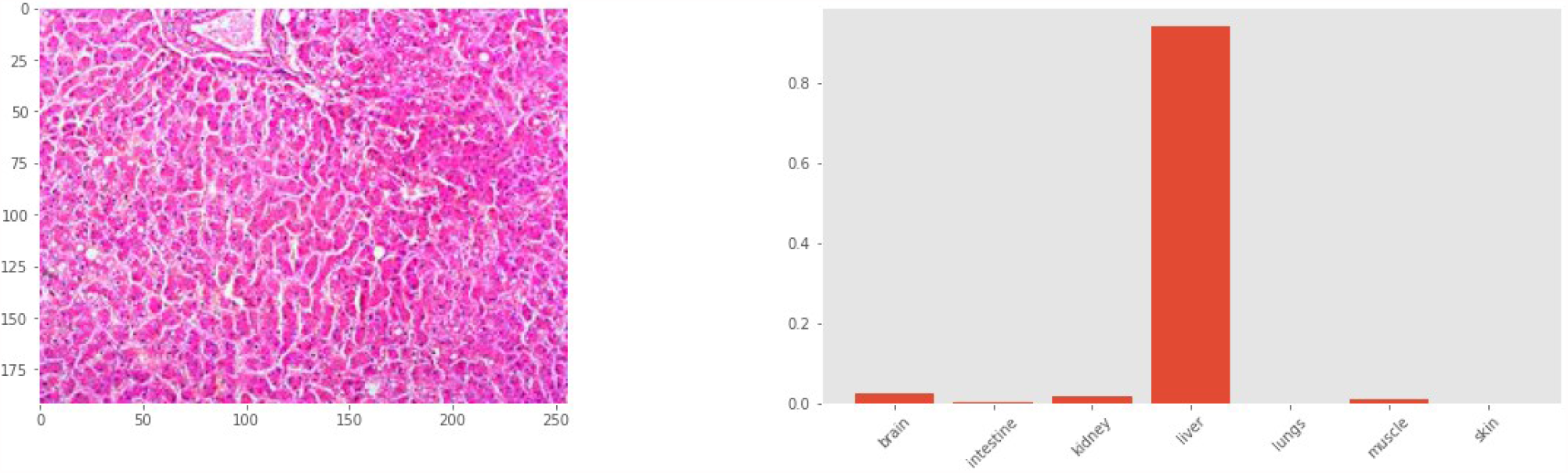
Liver correctly identified by the model

## Discussion

The analysis of histologic images is a non trivial machine learning problem, because of the inherent variability in biological tissues. No two foci from any tissue are exactly the same; even in a homogeneous organ like liver, one with well defined compact lobular architecture, arrangement of hepatocytes around central veins radiating towards portal triads show significant variability in each focus. Tis variability in architecture is present in all bodily tissues. For example, in the intestines, there might be significant variability in villus: crypt ratio along the length of the gastro intestinal tract.^[17]^ Similarly, depending on site of biopsy, histology of skin might show variable thickness of epidermis and stratum corneum. A learning model, either human or machine, must learn to take all the morphologic variability in consideration and learn the essential features of histomorphology which give tissues their identity.

The difficulties in histologic image analysis have been enumerated by Komura et al.^[18]^ One of the major difficulties encountered in Whole Slide Image (WSI) analysis is lack of labeled images. A pathologist must manually label a region of interest (ROI) an a WSI for the machine to train. We have not used whole slide images, but random foci from the slide to train the machine learning model. The entire image, not just a ROI, was used as input to the model. The other difficulty is that of magnification, because depending on magnification, the same tissue might show different histologic patterns. We have photographed all images at 10x magnification to have a consistent histologic pattern for the model to learn.

A similar study by Kieffer et al used grayscale histopathology images of 1000 x 1000 pixels, belonging to 24 classes, achieving 74.87% accuracy^[13]^ on 24 classes, using the feature vector from the last layer of a pretrained neural network. They concluded that the performance of a pretrained network and a network built specifically for the task were comparable. We have used color images and a pretrained CNN (VGG16), but altered its final, fully connected layers so that it produces only one of seven outputs. This model has achieved 88% accuracy, although on a smaller dataset than Kieffer et al.

Analysis of the predictions made by the model shows that the model has wrongly classified 14 images of kidney, predicting them as ‘liver’ (07), ‘lungs’ (03) or ‘muscle’ (04). This may be attributable to the deeply eosinophilic renal tubules in these images as well as overfitting on the ‘liver’ class. Kidney tissue is readily recognisable by human observers due to the prominent glomeruli, even if a single glomerulus is present. The fact that the model has often missed out on kidney tissue indicates a difference between how a human and a machine perceives a histologic image.

Analysis of the deeper layers of the model reveals a pattern. Figures 10-12 shows example images from the validation set with the first 3 slices of the first four layers of the CNN. The intermediate layers show the process of convolution, and how the features of the image are used to arrive at a simplified array of numbers. Figure 10 & 11, having at least 03 full or partial glomeruli, were classified correctly as kidney; whereas, figure 12 – having only one small glomerulus – was recognised as liver. Interestingly, the artifactual tear in the tissue (Figure 11) during sectioning has been lost over successive intermediate layers, indicating that the CNN is not affected by minor artifacts introduced during sectioning.

**Figure 5:**
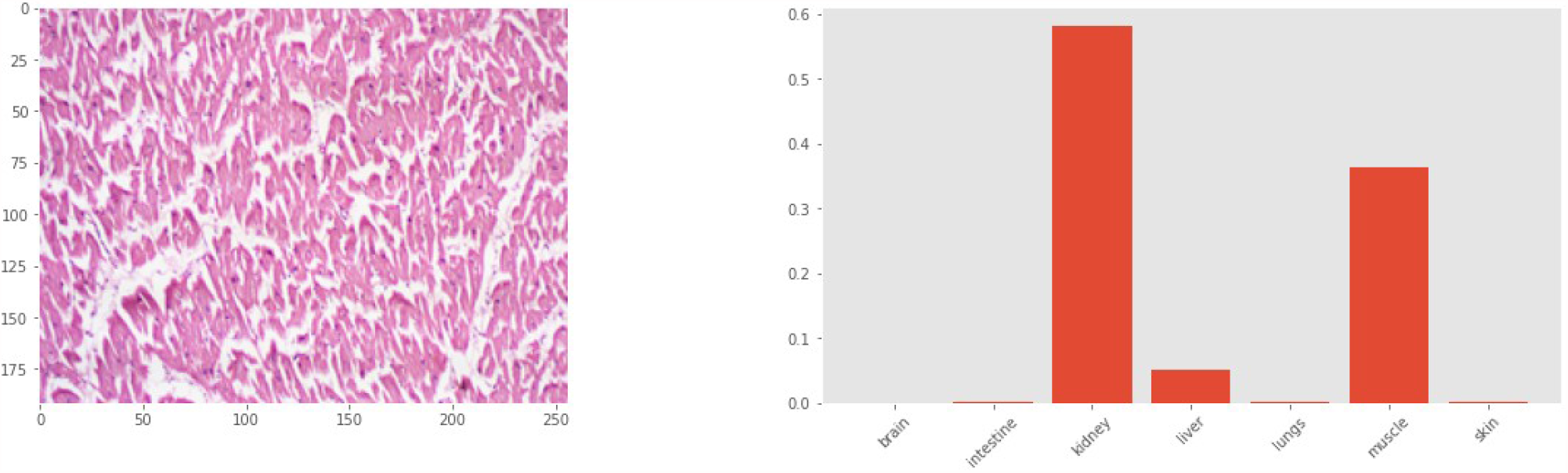
Muscle tissue wrongly classified by the model as kidney

**Figure 6:**
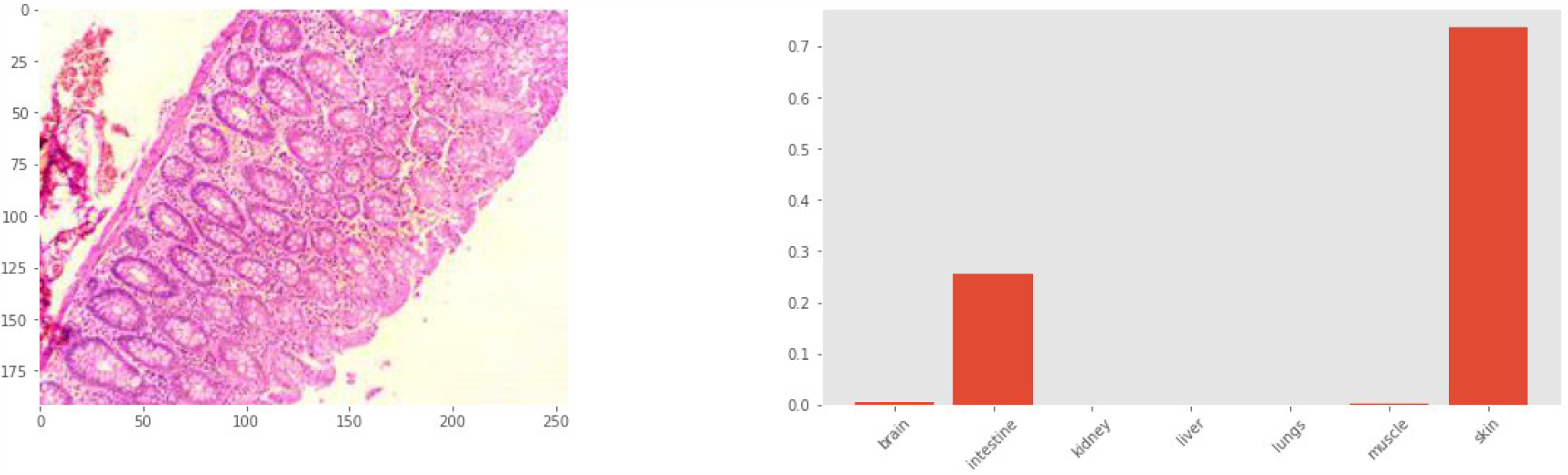
Intestine wrongly classified by the model; this might be due to the fact that the pattern of an epithelial layer overlying a fibrovascular stroma is common to both skin and intestine

**Figure 7:**
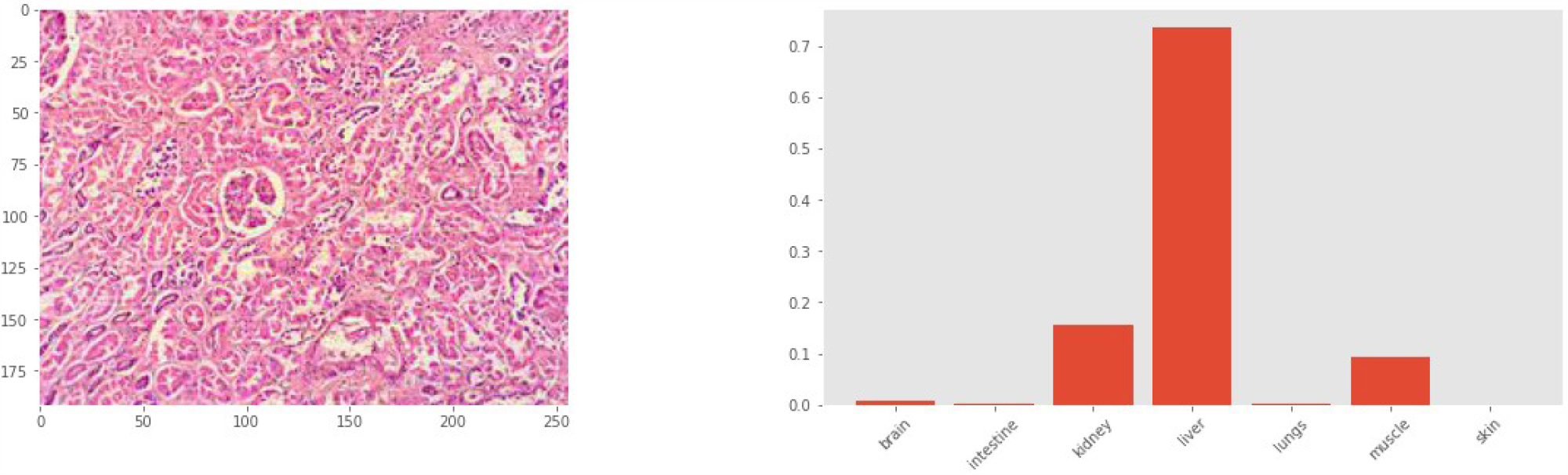
Kidney wrongly classified as liver; the single small glomerulus seems to have been ignored by the model

**Figure 8:**
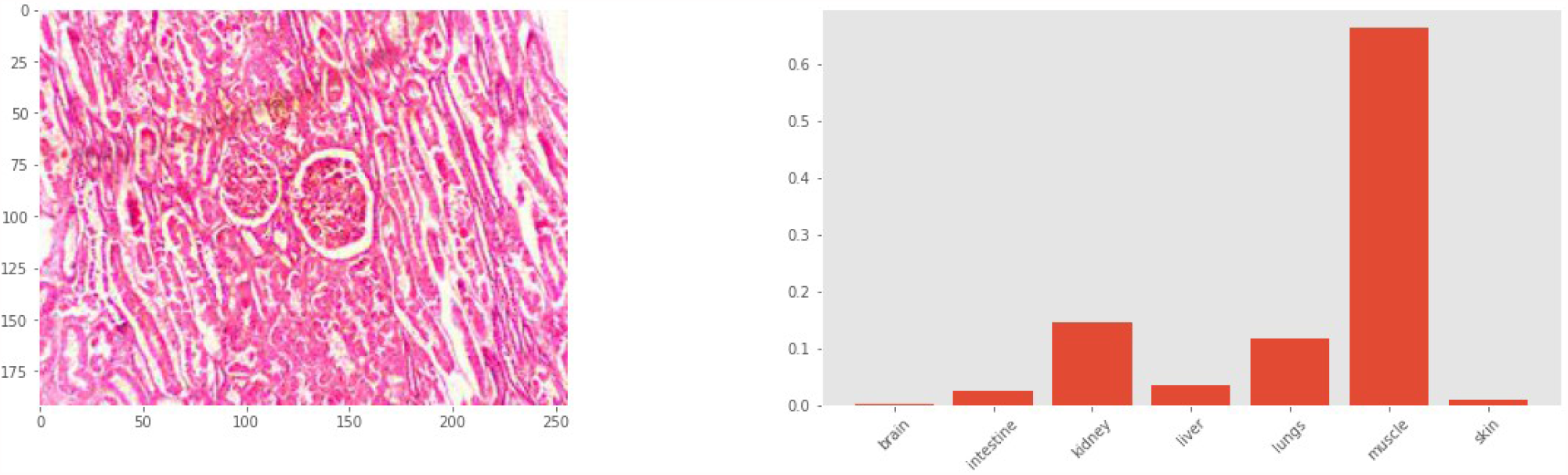
Kidney wrongly classified as muscle; the deeply eosinophilic staining produces a wrong impression of muscle tissue

**Figure 9:**
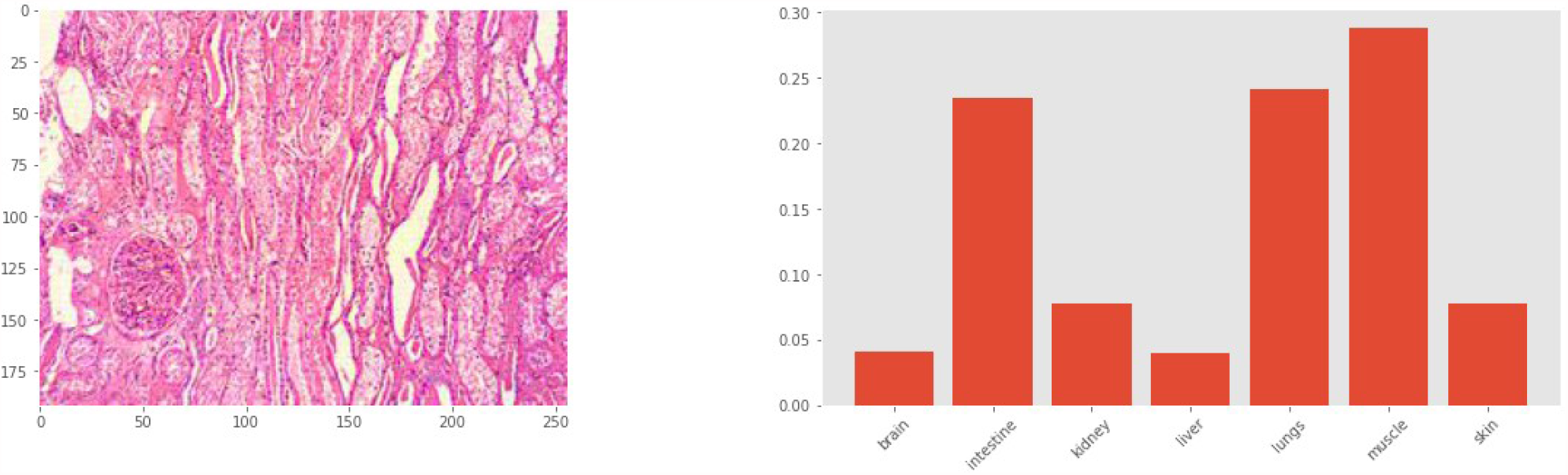
The model fails to classify this image (from kidney) in any definite category; there is deep eosinophilia resembling muscle, but also artifactual blank spaces – similar to alveoli in lungs

**Figure 10:**
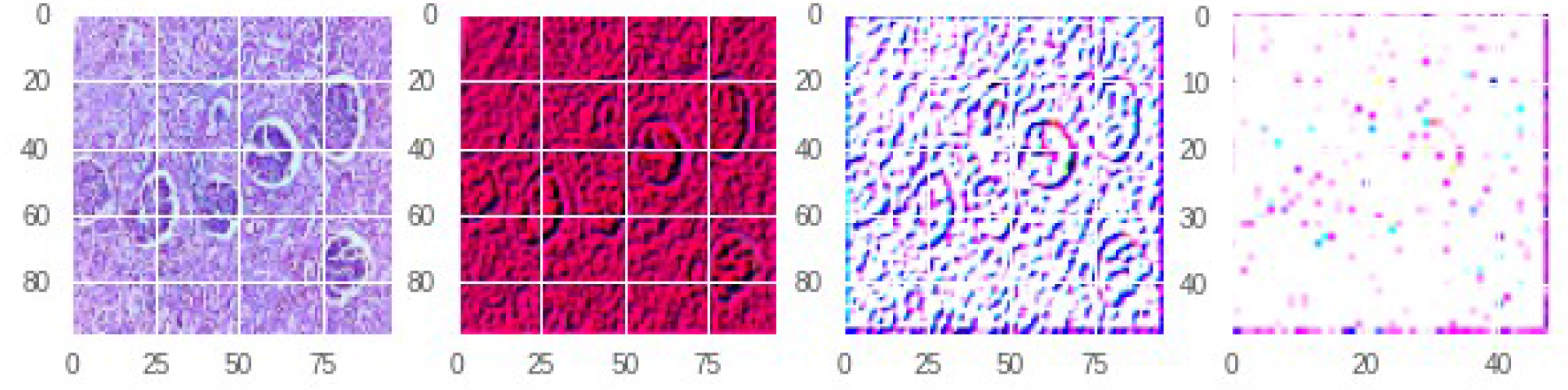
Inner layers of the network while correctly classifying an image from kidney; the glomeruli are well preserved till the third layer

**Figure 11:**
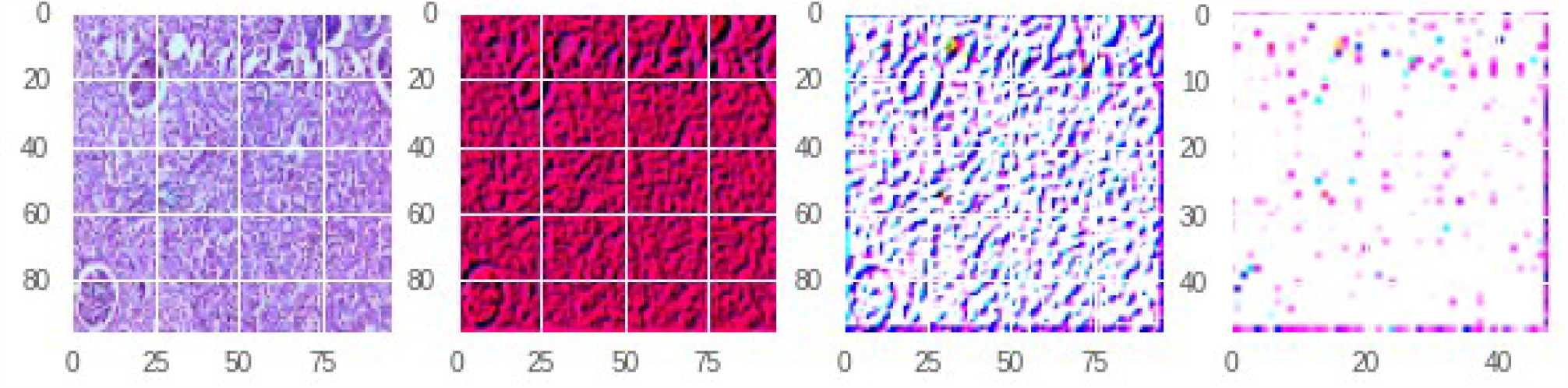
Another image from kidney, correctly classified, with activations in inner layers; note the presence of glomerular structures in the third layer

**Figure 12:**
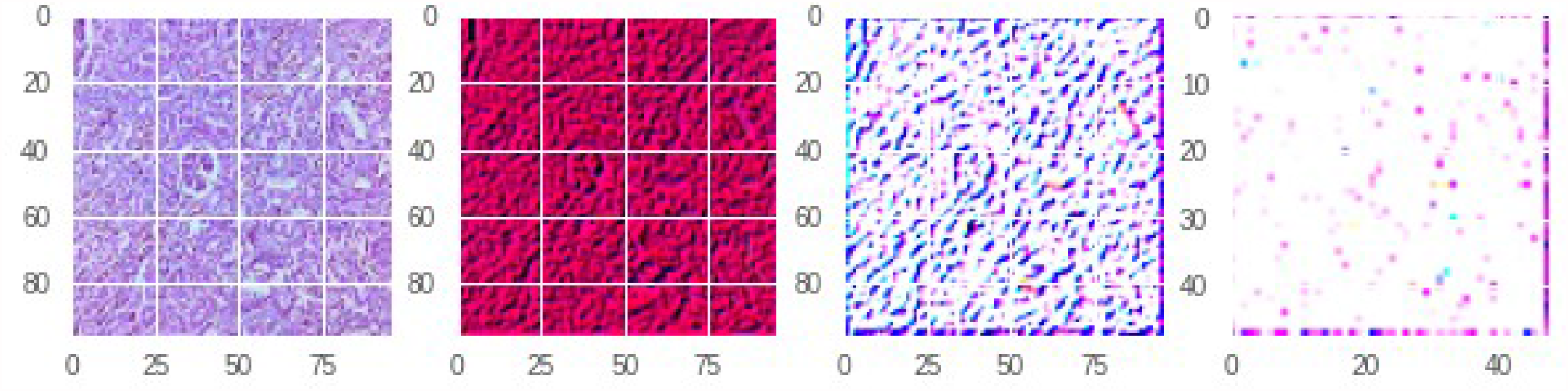
Kidney wrongly classified as liver, with activations in inner layers; the single small glomerulus is not preserved till the third layer

Figure 14 shows an image (muscle) which was wrongly classified by the CNN as ‘intestine’. The deeper layers of the CNN, while operating in this image, produce a characteristic pattern of smooth muscle layering around intestinal epithelia, which might be the cause of this error. Again, in figure 15, the artifactual blank spaces in the image has lead to the wrong classification as ‘lung’

**Figure 13:**
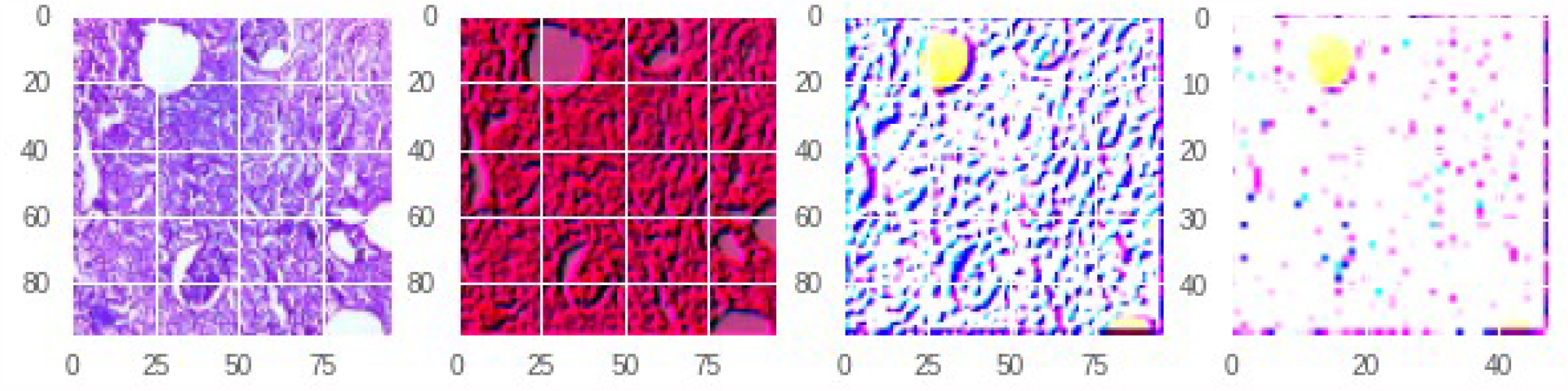
Kidney wrongly classified as lung; possibly due to the artifactual blank space (may have been mistaken for an alveolus)

**Figure 14:**
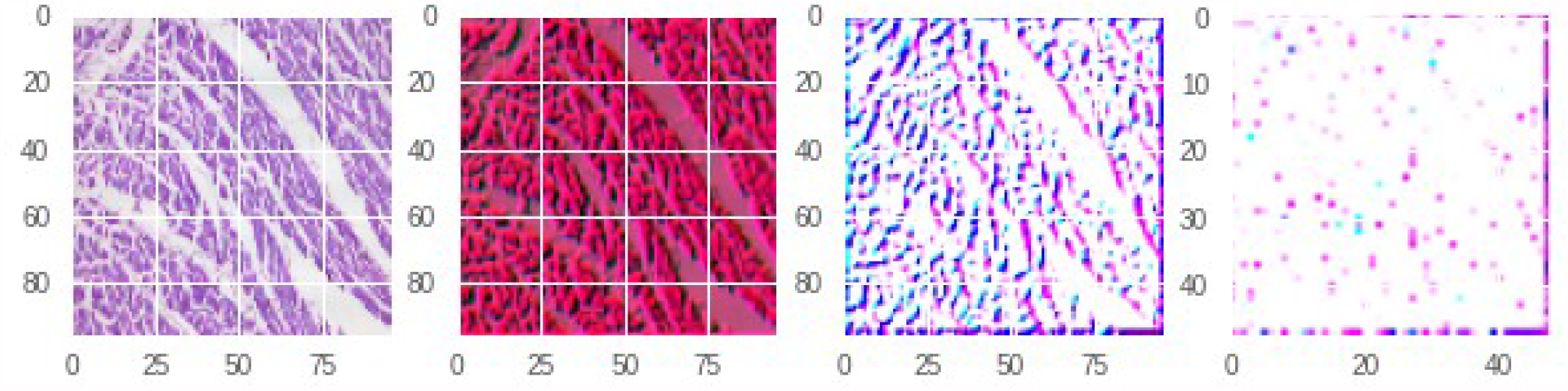
Image from muscle tissue wrongly classified as intestine; the layered pattern of muscle fibers in this image is reminiscent of intestinal smooth muscles

**Figure 15:**
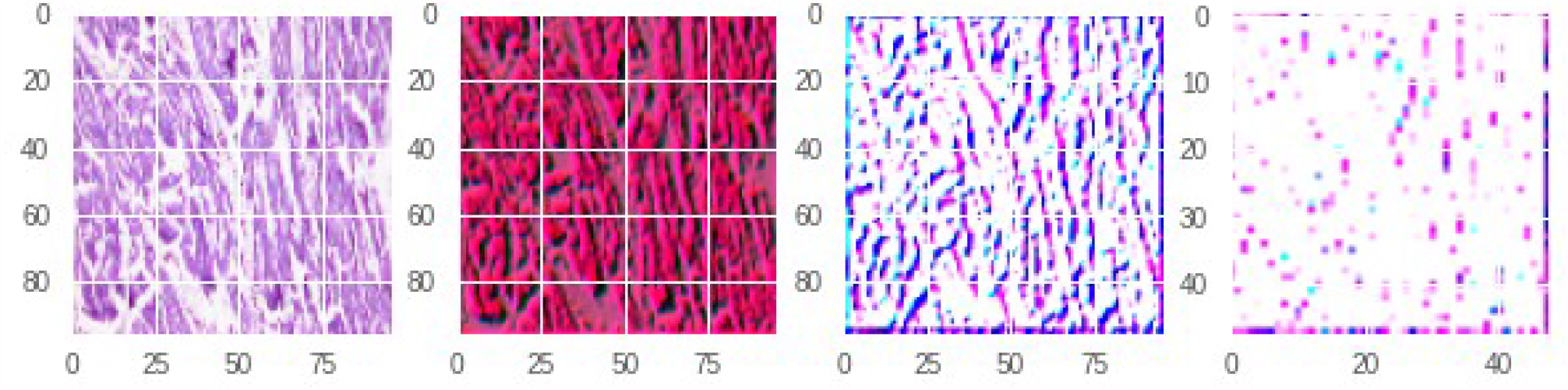
Muscle tissue wrongly classified as lung; note the abundant artifactual blank spaces in the original image

## Conclusion

The results of the present study indicates that a convolutional neural network might be trained in histology similar to a trainee pathologist, and is prone to similar kind of error as that of the beginner human pathologist. However, the study represents the first step towards developing a machine learning model as a generalised histopathological image classifier.

## Data Availability

Data and code available in public repository

https://github.com/cmacus/histo_jal

## Conflicts of interest

None to declare

## Note

Sayak Paul contributed to this work while employed at PyImageSearch

## Notes

### Competing Interest Statement

The authors have declared no competing interest.

### Funding Statement

No funding agency is involved

### Author Declarations

Institutional Ethical Committee MH Jalandhar (retrospective approval) 1. Dr Harish Chander Sharma, Registrar MH Jalandhar (retd) 2. Dr Rajesh Khanna, Head of the Dept of Surgery, MH Jalandhar 3. Dr Samrat Mitra, Dept of General Medicine, MH Jalandhar DECISION: APPROVED

## References

1. Krizhevsky A, Sutskever I, Hinton GE. ImageNet Classification with Deep Convolutional Neural Networks [Internet]. In: Proceedings of the 25th International Conference on Neural Information Processing Systems - Volume 1. USA: Curran Associates Inc.; 2012 [cited 2019 Jan 10]. page 1097–1105.Available from: http://dl.acm.org/citation.cfm?id=2999134.2999257

2. Simonyan K, Zisserman A. Very Deep Convolutional Networks for Large-Scale Image Recognition. ArXiv14091556 Cs [Internet] 2014 [cited 2019 Feb 13];Available from: http://arxiv.org/abs/1409.1556

3. CS386n Convolutional Neural Networks for Visual Recognition [Internet]. [cited 2019 Jan 10];Available from: http://cs386n.github.io/convolutional-networks/

4. Wei JW, Tafe LJ, Linnik YA, Vaickus LJ, Tomita N, Hassanpour S. Pathologist-level classification of histologic patterns on resected lung adenocarcinoma slides with deep neural networks. Sci Rep 2019;9(1):3358.

5. MRF-ANN: a machine learning approach for automated ER scoring of breast cancer immunohistochemical images. - PubMed - NCBI [Internet]. [cited 2019 Jul 17];Available from: https://www.ncbi.nlm.nih.gov/pubmed/28319275

6. Exploring automatic prostate histopathology image Gleason grading via local structure modeling. - PubMed - NCBI [Internet]. [cited 2019 Jul 17];Available from: https://www.ncbi.nlm.nih.gov/pubmed/26736836

7. Gertych A, Ing N, Ma Z, Fuchs TJ, Salman S, Mohanty S, et al. Machine learning approaches to analyze histological images of tissues from radical prostatectomies. Comput Med Imaging Graph Off J Comput Med Imaging Soc 2015;46(Pt 2):197–208.

8. Mitosis detection in breast cancer histological images An ICPR 2012 contest. - PubMed - NCBI [Internet]. [cited 2019 Jul 17];Available from: https://www.ncbi.nlm.nih.gov/pubmed/23858383

9. Quantification of tumour budding, lymphatic vessel density and invasion through image analysis in colorectal cancer. - PubMed - NCBI [Internet]. [cited 2019 Jul 17];Available from: https://www.ncbi.nlm.nih.gov/pubmed/24885583

10. Deep learning assessment of tumor proliferation in breast cancer histological images - IEEE Conference Publication [Internet]. [cited 2019 Jul 17];Available from: https://ieeexplore.ieee.org/abstract/document/8217719

11. Wang D, Khosla A, Gargeya R, Irshad H, Beck AH. Deep Learning for Identifying Metastatic Breast Cancer. ArXiv160605718 Cs Q-Bio [Internet] 2016 [cited 2019 Jul 17];Available from: http://arxiv.org/abs/1606.05718

12. Chen H, Qi X, Yu L, Heng P-A. DCAN: Deep Contour-Aware Networks for Accurate Gland Segmentation [Internet]. 2016 [cited 2019 Jul 17]. page 2487–96.Available from: https://www.cv-foundation.org/openaccess/content_cvpr_2016/html/Chen_DCAN_Deep_Contour-Aware_CVPR_2016_paper.html

13. Kieffer B, Babaie M, Kalra S, Tizhoosh HR. Convolutional Neural Networks for Histopathology Image Classification: Training vs. Using Pre-Trained Networks. ArXiv171005726 Cs [Internet] 2017 [cited 2019 Jul 17];Available from: http://arxiv.org/abs/1710.05726

14. Welcome to Python.org [Internet]. [cited 2019 Jul 17];Available from: https://www.python.org/

15. Keras [internet]; available from keras.io

16. Google Colaboratory [Internet]. [cited 2019 Jun 5];Available from: https://colab.research.google.com/notebooks/basic_features_overview.ipynb

17. Marsh MN, Rostami K. What Is A Normal Intestinal Mucosa? Gastroenterology 2016;151(5):784–8.

18. Komura D, Ishikawa S. Machine Learning Methods for Histopathological Image Analysis. Comput Struct Biotechnol J 2018;16:34–42.

